# Economic evaluation of a cluster randomized, non-inferiority trial of differentiated service delivery models of HIV treatment in Zimbabwe

**DOI:** 10.1101/2021.11.17.21266474

**Authors:** Mariet Benade, Brooke Nichols, Geoffrey Fatti, Salome Kuchukhidze, Kudakwashe Takarinda, Nicoletta Mabhena-Ngorima, Ashraf Grimwood, Sydney Rosen

## Abstract

**Background:** About 85% of Zimbabwe’s >1.4 million people living with HIV are on antiretroviral treatment (ART). Further expansion of its treatment program will require more efficient use of existing resources. Two promising strategies for reducing resource utilization per patient are multi-month medication dispensing and community-based service delivery. We evaluated the costs to providers and patients of community-based, multi-month ART delivery models in Zimbabwe.

**Methods:** We used resource and outcome data from a cluster-randomized non-inferiority trial of three differentiated service delivery (DSD) models targeted to patients stable on ART: 3-month facility-based care (3MF), community ART refill groups (CAGs) with 3-month dispensing (3MC), and CAGs with 6-month dispensing (6MC). Using local unit costs, we estimated the annual cost in 2020 USD of providing HIV treatment per patient from the provider and patient perspectives.

**Results:** In the trial, retention at 12 months was 93.0% in the 3MF, 94.8% in the 3MC, and 95.5% in the 6MC arms. The total average annual cost of HIV treatment per patient was $187 (standard deviation $39), $178 ($30), and $167 ($39) in each of the three arms, respectively. The annual cost/patient was dominated by ART medications (79% in 3MF, 87% in 3MC; 92% in 6MC), followed by facility visits (12%, 5%, 5%, respectively) and viral load (8%, 8%, 2%, respectively). When costs were stratified by district, DSD models cost slightly less, with 6MC the least expensive in all districts. Savings were driven by differences in the number of facility visits made/year, as expected, and low uptake of annual viral load tests in the 6-month arm. The total annual cost to patients to obtain HIV care was $10.03 ($2) in the 3MF arm, $5.12 ($0.41) in the 3MC arm, and $4.40 ($0.39) in the 6MF arm.

**Conclusions:** For stable ART patients in Zimbabwe, 3- and 6-month community-based multi-month dispensing models cost less for both providers and patients than 3-month facility-based care and had non-inferior outcomes.

## INTRODUCTION

Zimbabwe, a country in southern Africa of just under 15 million people, has a high burden of HIV infection, with a prevalence of more than 10% among adults.^1,2^ Great progress has been made in expanding access to testing and treatment, care, with an estimated 78% of all people living with HIV (PLHIV) on antiretroviral therapy (ART).^2^ Achieving the UNAIDS 95-95-95 targets, however, will require both further expansion of the treatment program and more efficient use of existing resources, as global donor support for HIV programs has plateaued.^3,4^

One promising strategy for expanding access to HIV treatment, improving outcomes, and increasing efficiency is the implementation and scale-up of differentiated service delivery (DSD) models. DSD models aim to move away from a “one-size-fits-all” approach to HIV service delivery and tailor care delivery to patient needs.^5^ DSD models typically adjust the location and frequency of service delivery, as well as the cadres of healthcare staff involved and specific services provided. ^6^ Examples include, but are not limited to, community adherence groups (CAGs), community-based medication pickup points, and multi-month dispensing (MMD) of medications.

In addition to reducing the burden of care on patients and achieving (at least) non-inferior clinical outcomes DSD models are expected to reduce costs for both patients and providers.^7–9^ A small number of recent studies from southern Africa suggest that some but not all DSD models allow for modest provider cost savings; some models have been found to increase provider costs.^10^ Nearly all DSD models studied have substantially reduced costs to patients, but by varying amounts.^7,11^ To our knowledge, no estimates of actual costs of DSD models in Zimbabwe have been published. Using data from a cluster randomized trial, we evaluated the costs to providers and patients of community-based, multi-month ART delivery models in Zimbabwe.

## METHODS

### Study setting and interventions

We used resource and outcome data from a cluster randomized, non-inferiority trial of DSD models for patients in Zimbabwe who met criteria as being stable on ART. The trial has previously been described in detail^12^. ART is provided free of charge to eligible residents. Patients were considered stable if they had an undetectable viral load and had remained on the same treatment regimen for at least 6 months. A total of 30 healthcare facilities (clusters) were randomized to offer one of three DSD models: three-month dispensing at the facility (3MF); three-month dispensing through a community ART refill group (3MC); or six-month dispensing through a community ART refill group (6MC). 3MF was an enhanced version of the standard of care at the time of the study in which patients visited a health facility every 3 months and received both a clinical consultation and a 3-month supply of ART at every visit. The 3MF arm was thus more resource intensive than actual standard of care, which required at least annual consultations only. The 3MC and 6MC arms utilized community ART refill groups (CAGs) of 6-12 patients each. A rotating patient representative of the CAG visited the facility and collected medications for all patients in the group every 3 or 6 months and then distributed medications to other group members at quarterly community meetings. All patients enrolled in CAGs were required to make a routine clinic visit for a consultation every 12 months.

### Outcomes

The primary study outcome was retention in care at 12 months, defined as 1-attrition. Attrition included loss to follow-up and all-cause mortality; loss to follow-up was defined as not collecting ART for ≥90 days during the follow-up period.^12^ For this economic analysis, we calculated: 1) the annual cost to the provider (healthcare system) per patient treated and per patient retained; and 2) the annual cost to access care from the patient perspective.

### Provider cost data collection and analysis

For the analysis from the provider perspective, we followed the same methodology as reported in a sister study conducted in Lesotho.^11^ We collected resource utilisation (visits, ART dispensed and laboratory tests performed) from patient records over the study period. Our data collection tool did not allow us to discern between scheduled facility and scheduled CAG visits. To calculate the number of visits by type, we assumed that all scheduled visits in the conventional care arm (3MF) were facility based. For the CAG arms, we assumed that two and one of the visits were facility visits for 3MC and 6MC, respectively.

### Unit cost estimates

Costs are reported in 2020 USD. Provider costs included the ART provided, laboratory costs, facility costs and the cost of delivering a CAG interaction.^13^ The cost of visits were calculated as the average cost of delivering a facility visit and CAG interaction as found in microcosting studies in two other countries.^10,11^ Equipment costs were annualized at a discount rate of 5% over their useful life years.

### Patient cost data collection and analysis

For the costs incurred by patients, we considered the transport costs to and from facility and CAG interactions, wages lost due to time seeking care and the total time patients spent accessing care. Data on patient costs were collected from a subset of 1,082 participants in all 3 study arms through semi-structured interviews with an 11-item questionnaire at study enrolment and the end of the 12-month follow-up period. Patients were asked about the number of visits to DSD interactions, the time spent on travel to access care, travel cost and distance travelled.

We assigned a value of lost wages to the opportunity costs of accessing care either at a facility or a CAG visit. To do this, we assumed an average daily wage of daily GDP per capita, which amounted to $1.50 per day, because Zimbabwe does not have a stated minimum wage.^14^ We assigned a full day’s wages lost for each facility visit and a quarter of a day’s wages lost for each CAG meeting. This assumption was based on interviews with DSD implementors and includes travel and waiting time for each DSD interaction.^11^

## RESULTS

The trial enrolled 4,800 participants (1,919 in 3MF, 1,335 in 3MC, and 1,546 in 6MC) (Table Supp 1). A detailed description of participants is provided elsewhere.^12^ Of relevance to this economic evaluation, most participants attended facilities in rural settings (80% in the 3MF arm, 71% in the 3MC arm, and 78% in the 6MC arm). Employment differed significantly between arms; 52% of 6MC, 40% of 3MF, and 24% of 3MC patients were employed. Most participants in all arms lived within 9 km of the nearest healthcare facility, but a significant minority (27% in 3MF, 18% in both 3MC and 6MC) lived 9 km or more from the nearest facility.

The trial demonstrated non-inferiority of outcomes in the two community CAG arms, compared to the facility-based arm. Retention in care at 12 months was highest in the 6MC arm (95.5%), followed by 3MC (94.8%) and 3MF (93.0%).

### Resource utilization and unit costs

Table 1 (panel a) presents average resource utilization per patient by study arm. The numbers of interactions with the healthcare system per patient per year were roughly consistent with model design: 5.01 visits per year in 3MF, 5.02 in 3MC, and 3.13 in 6MC. All five 3MF interactions were visits to the healthcare facility, while only two were facility visits in the 3 and 6-monthly CAG arms. CAG interactions (group meetings) in the two community-based arms differed, with a mean of 3.05/patient/year in 3MC and 1.18/patient/year in 6MC.

**Table 1.**
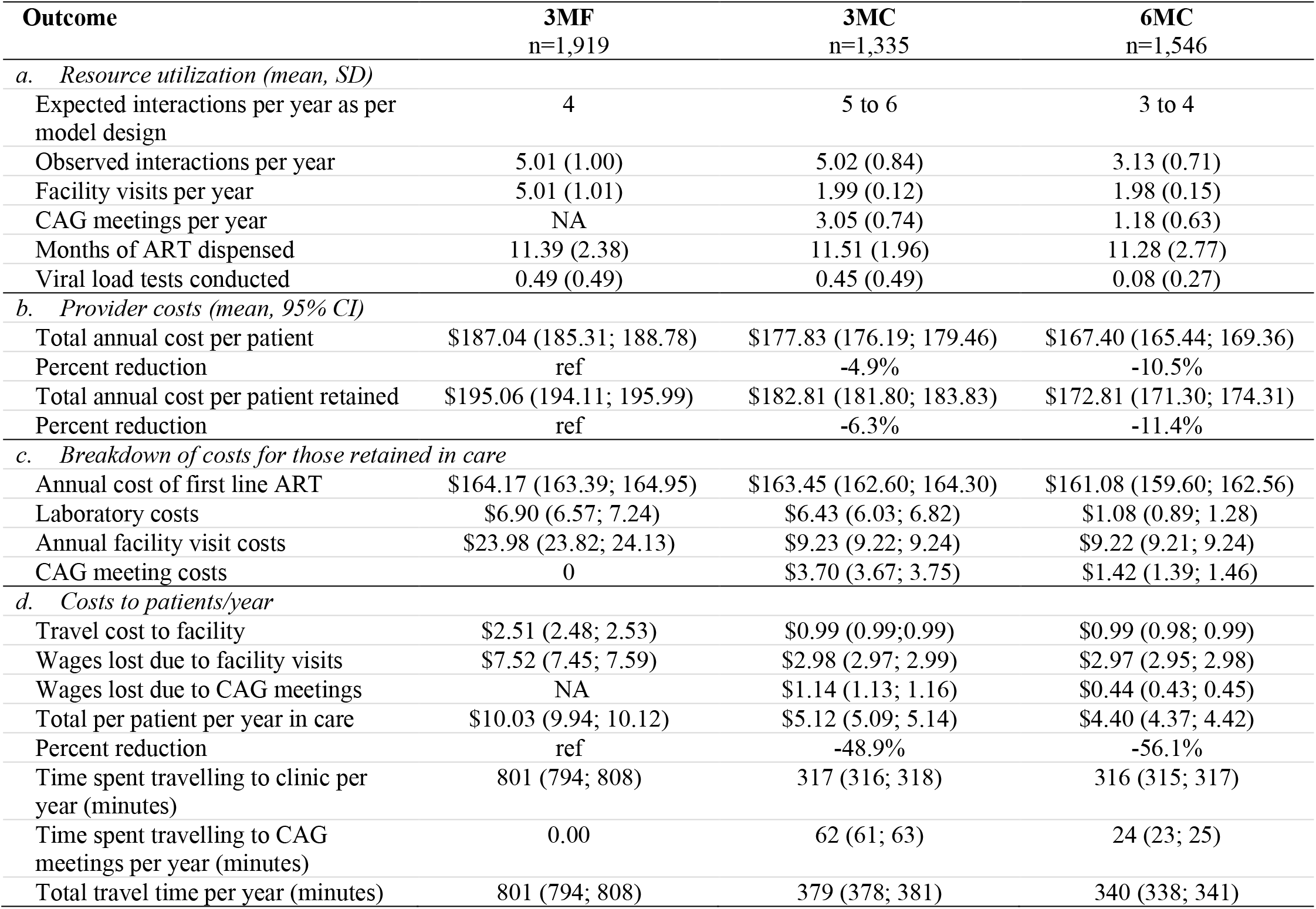
Average resource utilization, cost per patient per year and per patient retained, and cost to patients.

Months of ART dispensed did not differ significantly among any of the arms, with participants receiving 11.39 months in 3MF, 11.51 in 3MC, and 11.28 in 6MC. The number of viral load tests conducted— intended to average 1 per year—was very low in 6MC, with only 0.08 tests done per patient over the 12-month study period. This number rose to 0.49 and 0.45 in the 3MC and 3MF arms, respectively.

We estimated the all-inclusive cost of a facility visit as $4.62 and of a CAG interaction as $1.88. From existing literature, we assumed a cost of $14.42 per viral load test and $13.81 for a month of first-line ART.^15^

### Provider costs per patient

As shown in Table 1 (panel b), the mean annual cost to the provider per patient enrolled in the study was highest in the 3MF arm at $187.04 (95% CI $185.31-$188.78), followed by 3MC at $177.83 ($181.80-183.83) and 6MC at $167.40 (CI $165.44-$169.36). The same pattern held for those retained in care, with the 3MF arm costing $195.06 (95% CI $194.11-$195.99) per patient retained compared to $182.81 ($181.80-$183.83) in the 3MC arm and $172.81 ($171.30-$174.31) in the 6MC arm. Differences in costs (Table 1 panel c) between the facility-based arm (3MF) and the two community-based arms were driven by differences in visit costs--$23.98/patient/year in the 3MF arm compared to $9.23 and $9.22 in the 3MC and 6MC arms respectively. The difference in cost between the 3MF and 6 MF models reflects both the smaller number of CAG meetings in the 6MC model and, importantly, the small proportion of patients in 6MC who completed viral load tests during the year-long follow up period. This is further explored in the district-level analysis.

### Costs to patients

The average annual cost to patients to receive ART care was $10.03 (95% CI $9.94-$10.12) in the 3MF arm, $5.12 ($5.09-$5.14) in the 3MC arm and $4.40 ($4.37-4.42) in the 6MC arm (Table 1, panel d). Patient costs were driven by wages lost in attending facility visits, as each visit was associated with losing an entire day’s wages, while CAG meetings only resulted in a quarter of a day’s wages lost. Travel time followed a similar pattern: participants in the 3MF arm averaged 801 minutes (95% CI 794-808) of travel time for healthcare interactions per year, while those in the 3MC and 6MC arms averaged only 379 (378-381) and 340 (338-341) minutes, respectively.

### Variability among districts

To understand variability in provider costs by location, we also estimated average provider cost/patient retained in care by geographic district. Costs varied among districts by roughly 10% in the 3MC arm, 7% in the 3MF arm; and 3% in the 6MC arm (Table 2). Variability among districts was driven mainly by differences in viral load test utilization. The proportion of patients receiving a viral load test varied widely by geographic district, as well as by study arm, from a low of just 4% of patients in Beitridge District, which is primarily rural, in the 6MC arm to a high of 96% of patients in Chintingwiza District, which is primarily urban, in both study arms implemented there.^12^ Differences in viral load utilization stemmed in part from differential access to testing equipment—viral load testing capacity was being rolled out in Zimbabwe at the time of the study and had not yet reached all the study districts. Even in districts that appear to have had viral load testing capacity, however, such as Beitbridge, patients in community models were generally less likely to have tests than were patients in the facility-based model.

**Table 2.** District level viral load utilization and provider costs.

| District | Gutu<br>(n=535) | Zaka<br>(n=1,017) | Mberengwa<br>(n=1,661) | Beitbridge<br>(n=813) | Chitungwiza<br>(n=774) |
| --- | --- | --- | --- | --- | --- |
| **Proportion of participants receiving a viral load test during the 12-month follow up period (n, %)** |  |  |  |  |  |
| 3MF | 64 (20) | 134 (25) | 151 (43) | 164 (88) | 352 (96) |
| 3MC | 13 (17) | 131 (55) | 26 (7) | 34 (18) | 362 (96) |
| 6MC | 11 (17) | 43 (24) | 41 (5) | 18 (4) | -- |
| **Average provider cost per patient retained** |  |  |  |  |  |
| 3MF | \$188.97 (1.6; 330) | \$192.31 (0.7; 546) | \$194.92 (0.8; 347) | \$202.98 (1.0; 188) | \$200.59 (1.1; 373) |
| 3MC | \$182.71 (0.8; 75) | \$186.40 (0.5; 238) | \$175.79 (0.9; 381) | \$174.45 (2.1; 193) | \$192.03(0.6; 378) |
| 6MC | \$177.43 (2.4; 66) | \$ 175.98 (1.3; 179) | \$171.75 (0.7; 816) | \$172.80 (2.2; 416) | -- |

## DISCUSSION

In this economic evaluation of three differentiated models of ART delivery in Zimbabwe, we found that both 3- and 6-monthly dispensing through community ART groups reduced costs for both providers and patients, primarily by reducing the number of visits required to a healthcare facility. For patients who meet eligibility criteria—those who are regarded as “stable” on ART—the two community models also achieved non-inferior retention in care, as previously reported.^12^ They can thus both be regarded as cost-effective alternatives to facility-based care with 3 month dispensing.

In interpreting this finding of cost-effectiveness, a number of provisos should be kept in mind. First, cost savings per patient in the community models were relatively modest (5-10% less than the facility model), and, importantly, these savings will only be realized if resource allocation at facilities is also adjusted. Fewer patient visits to a clinic, for example, will only reduce healthcare budgets if the change allows the facility to pay for fewer human and physical resources. In most cases, clinics do not have the ability to match their physical space, staff complement, or other resources to relatively modest changes in demand for services, at least in the short term. Instead, the “savings” from enrolling patients in less intensive models of care—here defined to mean those requiring fewer clinic visits—are likely to be seen in shorter waiting times for patients, higher quality of care from clinicians, more time available for activities including record-keeping and outreach, and/or more time for staff breaks or shorter working hours. These changes (with the possible exception of the last one) may be of great value to the health system and its patients, but they will not result in monetary savings.

A second consideration is that one reason for the lower cost of the 6MC model, in particular, was the poor uptake of viral load testing in this arm. We believe the reasons to be a combination of geographic access—some districts did not yet have viral load testing capacity, and 6MC was overrepresented in these districts—and the small number of clinic visits made by 6MC patients. While we cannot be certain of the latter reason, the fact that two districts, Mberengwa and Beitridge, tested far more 3MF patients than 3MC or 6MC patients may indicate a need for more investment in compliance with annual viral load testing guidelines if patients receive services in the community, rather than at a facility.

And third, while the trial and this analysis used the 3MF arm to represent standard of care in Zimbabwe, the 3MF model in fact provided more care than currently is recommended by national guidelines in Zimbabwe, which require only one clinical consultation a year. We thus cannot be sure that true standard of care costs as much as the 3MF model analyzed here, though our study design accounts for WHO guidelines that recommend clinical visits 3-6monthly for patients stable on ART.^16^

Our findings that DSD models are cost saving to both the provider and patient while achieving similar results in retaining patients in care add to a small but growing evidence base that at least some DSD models cost less than conventional, facility-based care. A parallel study conducted in Lesotho reported a similar saving (6%) to providers and substantial savings to patients^11^, and a trial of facility-based MMD in Malawi and Zambia found that increasing the dispensing interval can lead to small cost savings even if all interactions occur at a health facility.^7,17^ A study of a variety of DSD models in Zambia, however, found that the models considered were slightly more expensive than conventional care, probably because the models did not reduce the resources required per patient treated. In all studies that have measured them, savings to patients in time and money tend to be large, an important consideration even if providers’ budgets are not affected.^7,18^

In addition to the considerations above, our study had two other limitations. First, the total number of interactions per participant was captured in the study, but not the type of visit. The breakdown of visits was therefore based on the model as recommended. Second, we did not collect patient cost data on all patients and therefore had to assign costs on a per-visit basis by visit type, based on average costs reported by the subset of participants who completed the patient cost questionnaire.

Despite these limitations, this study demonstrates that in Zimbabwe, community ART groups can save money for both providers and patients, particularly when combined with 6-month dispensing, without compromising patient outcomes. The consistency of this finding with those of other studies suggests that for stable, adult ART patients, offering community-based models of care makes sense for the healthcare system. The next challenge will be to identify cost-effective models of care for patients who do not meet current criteria for stability, such as a suppressed viral load or experience on ART, and for children, adolescents, and other groups.

## Data Availability

Requests for data will be considered together with a protocol approved by the Zimbabwe Medical Research Council and approval from the Zimbabwe Ministry of Health and Child Care.

## Acknowledgements

None

## Authors’ contributions

MB analyzed the data and drafted the manuscript. SR and BN designed the analysis and contributed to drafting the manuscript. SK contributed to data analysis. GF, KT, NMN, and AG designed and conducted the original study. All authors reviewed and approved the final manuscript.

## SUPPLEMENTARY MATERIAL

**Supplemental table 1.** Baseline characteristics

| **Characteristic (n (%))** | **3MF**<br>**(n=1,919)** | **3MC**<br>**(n=1,335)** | **6MC**<br>**(n=1,546)** |
| --- | --- | --- | --- |
| Age (median, IQR) | 45 (38-53) | 47 (41-56) | 45 (38-54) |
| Sex |  |  |  |
| Male | 541 (28) | 351 (26) | 445 (29) |
| Female | 1378 (72) | 984 (74) | 1101 (71) |
| Setting |  |  |  |
| Urban | 387 (20) | 387 (29) | 337 (22) |
| Rural | 1532 (80) | 948 (71) | 1209 (78) |
| Currently employed |  |  |  |
| Yes | 765 (40) | 319 (24) | 800 (52) |
| No | 1148 (60) | 1009 (76) | 741 (48) |
| Unknown | 6 (0.3) | 7 (1) | 5 (0.3) |
| Living >9km from facility |  |  |  |
| Yes | 520 (27) | 236 (18) | 273 (18) |
| No | 1388 (72) | 1098 (82) | 1272 (82) |
| Unknown | 11 (1) | 1 (0.0) | 1 (0.0) |
| Retained |  |  |  |
| Yes | 1784 (93) | 1265 (95) | 1477 (96) |
| No | 135 (7) | 70 (5) | 69 (4) |

